# Different Methods of Ankle-Brachial Index Calculation on the Prevalence of Peripheral Arterial Disease and Arterial Calcification in Subjects with Type 2 Diabetes Mellitus from a Public Hospital in Peru

**DOI:** 10.1101/2024.12.22.24319510

**Authors:** Luis Fernando Espinoza-Enciso, Iván Gonzalo Hernández-Gozar, Kevin Clared Zuñiga-Baldarrago, Robert Lozano-Purizaca, Manolo Briceño-Alvarado, Marlon Yovera-Aldana

**Affiliations:** School of Medicine, Universidad Científica del Sur, Lima, Perú; School of Medicine, Universidad Nacional de Piura, Piura, Perú; Grupo de Investigación MBA Cardiovasculares, Lima, Perú; Villamedic Groups, Lima, Perú; Grupo de Investigación de Neurociencias, Metabolismo y Efectividad clínica y Sanitaria, Universidad Científica del Sur, Lima, Perú

**Keywords:** Peripheral Arterial Disease, Arterial Calcification, Ankle-Brachial Index, Diabetes Mellitus, Prevalence, Risk Factors

## Abstract

**Objective:** To determine the frequency of peripheral arterial disease (PAD) and arterial calcification (AC) using three methods of ankle-brachial index (ABI) in patients with type 2 diabetes mellitus.

**Methodology:** A descriptive cross-sectional study using data from the Foot at Risk Program of the Endocrinology Department at Hospital María Auxiliadora from 2015 to 2020. We calculated the ABI for each lower limb using the lowest, highest, or average systolic pressure from the dorsalis pedis or posterior tibial artery in the ipsilateral leg. We defined PAD if any ABI from either leg was <0.9; AC if ABI was >1.3; and the rest were classified as normal. We calculated the prevalence of PAD and AC for each ABI method and assessed differences between ABI categories based on clinical variables using the chi-square or Fisher’s exact test.

**Results:** We included 643 subjects with a mean age of 61.4 years, 69.8% female. The prevalence of PAD was 7.8%, 15.4%, and 28.2% using the highest, average, or lowest systolic pressure as the numerator in the ABI, respectively. The highest prevalence of PAD occurred with the lowest pressure and the lowest with the highest pressure. AC was observed in 18.2%, 11%, and 16.2%, and normal values were 74%, 73.6%, and 55.7%. In all three methods, PAD was associated with older age and AC was associated with longer duration of diabetes.

**Conclusions:** Using different ABI methods, we observed a prevalence of PAD ranging from 7.8% to 28%, and a prevalence of CA between 11% and 18% among patients with type 2 diabetes mellitus at a public hospital in Peru. Further research is necessary to verify if the new ABI calculation methods provide improved accuracy in predicting complications.

## Introduction

Atherosclerotic diseases continue to progress globally due to non-communicable conditions such as diabetes mellitus, hypertension, and dyslipidemia, exacerbated by increased life expectancy and unhealthy lifestyles [1]. Currently, these diseases are the leading cause of death in high, middle, and low-income countries[2]. Among these conditions, peripheral arterial disease (PAD) is particularly associated with a high risk of lower limb amputation, especially in patients with diabetes mellitus (DM) [3]. In patients with DM, PAD typically presents as infrainguinal, bilateral, with medial layer calcifications and is often asymptomatic due to peripheral neuropathy [4]. These characteristics require considerations for efficient diagnosis, treatment, and follow-up management [5].

Globally, between 11.2% and 14.3% of patients with diabetes present some degree of PAD [6]. Critical ischemia or advanced PAD contributes to 30% of amputations, and 20% of these patients die within six months [7]. Reports from the Peruvian epidemiological surveillance system show low prevalences, suggesting significant underreporting due to a lack of adequate screening[8]. In patients hospitalized for diabetic foot in 36 Peruvian hospitals, the prevalence of PAD was 18.9, although there is much heterogeneity due to various methods used [9]. In high-income countries, there are no significant gender differences, and its prevalence increases with age (5% at 45-49 years and 18% at 85-89 years). Conversely, in low/middle-income countries, there is a predominance in women (2.7% to 6.3% in women and 1.2% to 3.9% in men)[10,11]. Other independent factors include a history of hypertension and dyslipidemia [12], as well as low levels of antioxidants [13].

Initial diagnostic methods are based on clinical scales such as Rutherford and Fontaine [14]. However, many patients with DM, due to painless neuropathy, may not present symptomatic indicators for diagnosis [15]. Among objective tests, the Ankle-Brachial Index (ABI) is one of the most accessible. This involves comparing the highest systolic blood pressure (SBP) at the ankle with the highest SBP at the arm. Generally, the accepted cutoff point for detecting PAD is 0.9 (5). Based on this point, a meta-analysis found that the ABI compared with angiography or Doppler ultrasound, presented an AUROC of 0.76. [16] Other systematic review found a low sensitivity of 63.5%, and a high specificity of 89.3% in patients with diabetes for the same test.[17] This reinforces the idea that, although the ABI is useful, it is not fully reliable for ruling out PAD in these patients. Therefore, other methods would be required to identify those high-risk patients without significant lesions. However, recent consensuses recommend methods that involve using the lowest systolic pressure at the ankle or the average of both arteries to detect less severe cases that might escape the traditional definition, while using the highest systolic pressure at the ankle would be aimed at determining the severity of the pathology [18].

Additionally, due to atherosclerosis, many arteries can be calcified, limiting compression and the calculation of the ABI. Therefore, other methods such as the Toe-Brachial Index, arterial plethysmography, or skin oximetry are necessary to accurately estimate vascular sufficiency [19]. Arterial calcification, being part of the atherosclerosis process, should not be overlooked in the context of cardiovascular disease [20].

Reports of PAD often exclude cases with arterial calcification, making it crucial to characterize this population given the inadequate metabolic control of patients [21]. Between 2% and 10% of patients in outpatient clinics of Peruvian hospitals present glucose, blood pressure, and lipids out of range[22,23]. Furthermore, the recommendation to adopt new criteria for calculating the ABI, including less severe cases, support the need to estimate the entire spectrum of possible outcomes [18]. Therefore, the objective of this study was to determine the frequency of peripheral arterial disease and arterial calcification using the ankle-brachial index in patients with type 2 diabetes mellitus at Hospital María Auxiliadora.

## Materials And Methods

### Study Design and Clinical Setting

An observational, cross-sectional, and descriptive study was conducted at Hospital María Auxiliadora, Lima, Peru, utilizing the database from risk foot consultations for patients with type 2 diabetes (DM2) in the endocrinology department, covering the period from December 1^st^ of 2015 to January 31th of 2020. This hospital is part of the Lima Sur Health Directorate.

### Population and Sample

The database included patients with all types of diabetes who had no issues performing the ankle-brachial index (ABI), which included not having lesions or wounds on lower limbs, severe pain in lower limbs that hindered the placement of the sphygmomanometer cuff, poorly controlled blood pressure, or severe edema in lower limbs. For this study, records with complete ABI data were included.

Records with four complete ABIs were included, taking the entire available population and employing a non-probabilistic, census sampling method.

### Variables

Ankle-Brachial Index (ABI): This is the result of a mathematical calculation between the systolic pressures of the lower and upper limbs and was estimated using three methods applied to the numerator of the index. The lowest, highest, and average systolic pressure from the ipsilateral pedal or posterior tibial artery were used as the numerator. The highest brachial pressure was used as the denominator. PAD was defined as any ABI <0.9; arterial calcification was defined as an ABI >1.3; and the rest were categorized as normal. [24] (**Fig 1**).

**Fig 1.**
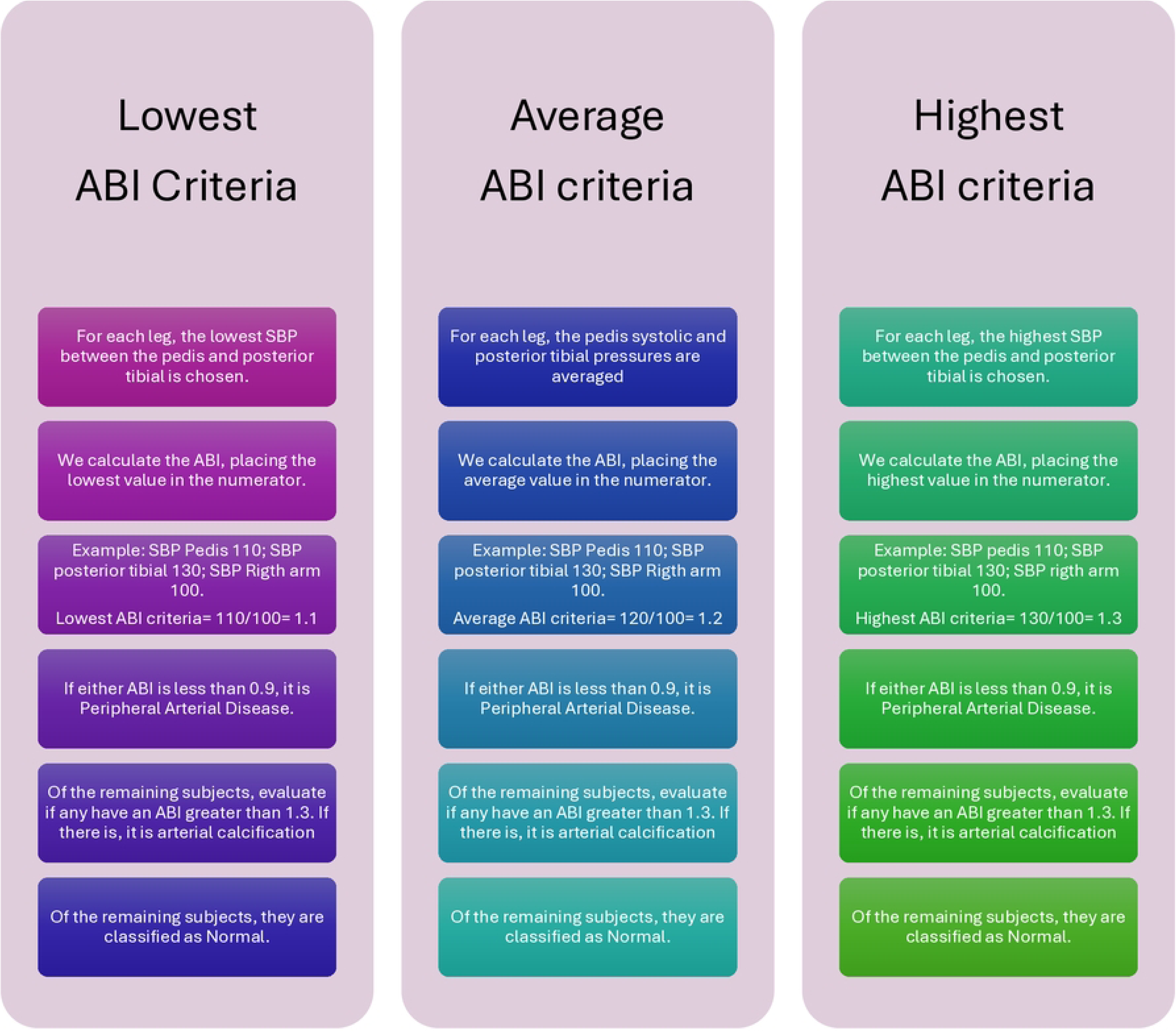
Diagnostic flowchart for PAD according to ABI criteria.

Other Variables: Demographic variables described included sex (male/female), age group (<60 years/61 to 74.9/75 and older), and education level (none/primary/secondary/higher). History variables included: stroke (no/yes), coronary artery disease (absent/present), hypertension (absent/present), and chronic kidney disease (absent/present). Diabetes-related variables included: disease duration (0-30 years), diabetic neuropathy (absent/present), and diabetic retinopathy (absent/present). Laboratory results included: hypercholesterolemia (less than 100 mg/dl/more than 100 mg/dl), hypertriglyceridemia (triglycerides less than 150 mg/dl/more than 150 mg/dl), glycated hemoglobin (less than 7%/7% or more). Poor metabolic control was defined as a glycated hemoglobin level equal to or greater than 7%, following the recommendations of the American Diabetes Association [25]. Systolic blood pressure was considered high if ≥140 mmHg and/or diastolic pressure ≥90 mmHg for individuals at low cardiovascular risk (less than 15% over 10 years). Concerning cholesterol levels, the goal was LDL < 100 mg/dl, in addition to HDL ≥ 40 in men or ≥ 50 in women and triglycerides ≥ 150 mg/dl. [26]

### At-Risk Foot Program

The At-Risk Foot Program of the Endocrinology Service at Hospital Nacional María Auxiliadora conducts clinical assessments based on the guidelines of the "International Working Group of Diabetic Foot Guideline on the Prevention of Foot Ulcers in Persons with Diabetes 2015." It assesses symptoms using the Total Symptom Score and foot care habits. It conducts foot inspections, assesses deformities, and evaluates protective, proprioceptive, and Achilles reflex sensitivity, while verifying pulse conditions. Systolic pressures in both arms and in the posterior tibial and pedal arteries were measured using a Yuwell mercury column sphygmomanometer and a Huntleigh pocket Doppler ultrasound device of 8 MHz.

The evaluation was carried out by endocrinologists with experience and training in diabetic foot care.

### Procedures

Permission was requested from the Head of the Endocrinology Service for authorization to use the data. The database was downloaded without names or personally identifiable information. Data were evaluated and analyzed from March 1 to June 30, 2024. The authors filtered records according to the eligibility criteria established. A cleaning of the database was performed to search for extreme data. No imputation was conducted in the case of missing data.

### Analysis Plan

The database was evaluated in Microsoft Excel to identify extreme values, incomplete data, or inconsistencies. Subsequently, STATA® V18 software (Stata Corp, College Station, Texas, USA) was utilized for data analysis.

The Ankle-Brachial Index (ABI) calculation was performed in a spreadsheet, following the algorithm in Figure 1. This process was carried out in the following order: first, patients with peripheral arterial disease (PAD) were identified, then those with arterial calcification among the remaining patients were selected, and finally, the unclassified cases were considered normal. These three categories were established for each of the three methods: highest, lowest and average ABI criterion. The prevalence of PAD, arterial calcification, and normal for each method was calculated along with its 95% confidence interval.

The absolute and relative frequencies of categorical variables were described. For quantitative variables, the mean and standard deviation were calculated if the data met the criteria for normality. Otherwise, the median and interquartile range were reported. The Shapiro-Wilk test was applied to determine the normality of the data.

A description of the ABI categorized into PAD, normal, and arterial calcification was performed according to demographic, clinical, and laboratory characteristics. This was done for each of the three methods of ABI calculation: highest pressure criterion, lowest pressure criterion, and average pressure criterion.

Using Poisson regression with robust variance, the crude prevalence ratio (PR) was calculated between cases of PAD and arterial calcification concerning the normal category for the variables of age, education level, duration of diabetes, sex, BMI, history of ulcer, peripheral neuropathy, and glycemic control. A multivariate analysis was conducted to obtain the adjusted prevalence ratios (aPR). Adjustments were made for variables that showed a p-value less than 0.2. Variables that were part of the definition of PAD (absence of pulse) were not adjusted. A significance level of 5% was used for all analyses.

We calculated the sample size power of 643 subjects to estimate the prevalence of peripheral arterial disease (PAD) for each criterion using the power command in STATA® V18. We employed the formula for estimating a proportion. We considered expected PAD prevalences based on Schröder’s study of 3.7%, 5.4%, and 14.6% for the highest, average, and lowest ABI criteria, respectively. [27] Additionally, we accounted for the proportions obtained in our study of 7.8%, 15.4%, and 28.2%. By configuring a two-tailed analysis and a 95% confidence level, we achieved powers of 99%, 100%, and 100%, respectively.

### Ethical Aspects

Approval was obtained from the Institutional Ethics and Research Committee of Hospital María Auxiliadora under code HMA/CIEI/015/2024. The data did not contain any personally identifiable information.

## RESULTS

From January 2015 to February 2020, a total of 1,019 patients were enrolled in the at-risk foot program database of María Auxiliadora Hospital. After excluding those with a history of major amputation (n=5), stroke (n=2), and subjects with incomplete ABIs (n=369), 643 subjects were included, who were evaluated using three methods for calculating the ABI: highest systolic pressure, average pressure, and lowest pressure. The results showed variations in the prevalence of PAD and arterial calcification (AC) according to the method of ABI calculation used.

**Figure 2.**
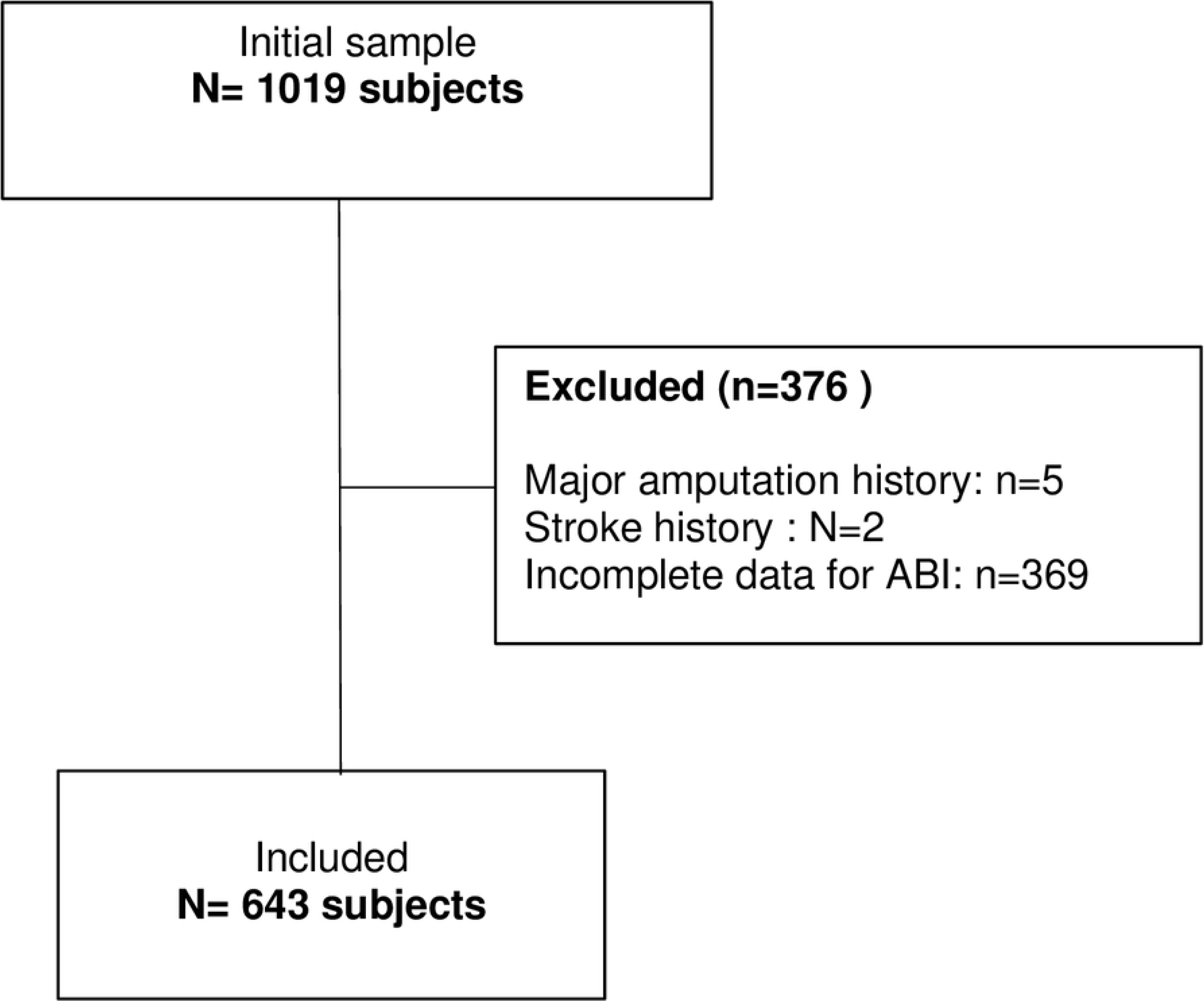
Flowchart of patients included in the study.

### General Characteristics

The sample presented a mean age of 61.4 years, of which 69.8% were women. Forty percent of the patients had more than 10 years since their diabetes diagnosis, and 31.9% used insulin alone or in combination with oral antidiabetic drugs as treatment. Among the most important comorbidities, 33.3% had hypertension, 8.6% had a history of ulcer, and 1.4% had a previous coronary artery disease.

Additionally, 28.9% presented painless peripheral neuropathy, while 61.4% experienced neuropathic pain. Regarding metabolic control targets, only 22.4% had a glycated hemoglobin level below 7%. When evaluating the integrated target of glycated hemoglobin lower than 7%, LDL below 100 mg/dl, and systolic blood pressure below 140 mmHg, only 4.4% met these criteria (**Table 1**).

**Table 1.** Demographic, Clinical, and Laboratory Characteristics of Patients with Diabetes Mellitus Treated at Hospital María Auxiliadora 2015-2020.

|  | Records | N (%) |
| --- | --- | --- |
| **Demographics** |  |  |
| Male, n(%) | 643 | 194 (30.2) |
| Age (years), Media SD | 643 | 61.4 (11.0) |
| Under 60 |  | 291 (43.7) |
| 60 to 74.9 |  | 276 (42.9) |
| 75 or older. |  | 76 ( 11.8) |
| Education level, n(%) | 643 |  |
| Illiterate |  | 39 (6.1) |
| Elementary |  | 219 (34.1) |
| High-School |  | 311 (48.4) |
| College |  | 74 (11.5) |
| **Patological history** |  |  |
| Diabetes diagnosis time (years), Median [IQR] | 643 | 7 [3 to 14] |
| Under 10 |  | 382 (59.4) |
| 10 to 19.9 |  | 180 (28.0 ) |
| 20 or more |  | 81 (12.6 ) |
| Diabetes medication, n(%) | 643 |  |
| Not medication |  | 34 (5.3) |
| OAD |  | 367 (57.1) |
| Inuline & OAD |  | 37 (5.8) |
| Only insuline |  | 205 (31.9) |
| Ulcer history | 643 | 55 (8.6) |
| Diabetic retinopathy history | 346 | 32 (9.3) |
| Treated hypothyroidism history | 598 | 2 (0.3) |
| Coronary disease history | 500 | 7 (1.4) |
| Hypertension | 408 | 136 (33.3) |
| **Clinical evaluation** |  |  |
| Neurological examination, n(%) |  |  |
| Alterad monofilament 10g <sup>a</sup> | 643 | 221 (34.7) |
| Altered tuning fork 128 Hz <sup>b</sup> | 643 | 243 (37.8) |
| Altered Achilles reflex <sup>c</sup> | 643 | 170 (26.4) |
| Peripheral neuropathy, n(%) |  | 186 (28.9) |
| Painful neuropathy, n(%) | 643 | 395 (61.4) |
| ABI n(%) | 643 |  |
| Some ABI < 0.9 |  | 167 (26.0) |
| Some ABI >1.3 |  | 104 (16.1) |
| Both |  | 14 (2.2) |
| Normal |  | 358 (55.7) |
| Decreased pedis or posterior tibial pulse | 643 | 155 (24.1) |
| BMI (kg/m<sup>2</sup>), median [IQR] | 425 | 27.3 [24.2 to 30.5] |
| SBP (mm Hg), median [IQR] | 643 | 137 [123 a 155] |
| **Laboratorio** |  |  |
| Glucose (mg/dl) , median [IQR] | 400 | 150 [117 to 196,5] |
| A1c (%),median [QIR] | 325 | 9.2 [7.1 a 11.7] |
| Creatinine (mg/dl), median [IQR] | 378 | 0.77 [0.63 to 0.98] |
| Estimated glomerular filtration rate < 60 ml/min, n(%) <sup>d</sup> | 379 | 85.2 (27.4) |
| Hemoglobin (g/dl), median [IQR] | 338 | 13.0 [11.2 a 14.6 ] |
| HDL-c (mg/dl), median [IQR] | 374 | 51 [40 a 67] |
| LDL-c (mg/dl), median [IQR] | 373 | 121 [89 a 151] |
| Triglycerides, median [IQR] | 357 | 151 [114 a 192] |
| Albuminuria (mg/24h), median [IQR] | 276 | 10.3 [5.9 a 15.6 ] |
| Albuminuria 30 mg/24 h, , n(%) |  | 33 (8.3) |
| **Metabolic control, n(%)** |  |  |
| A1c < 7 % | 325 | 73 (22.4) |
| LDL-c < 100 mg/dl (2.58 mmol/L | 373 | 140 (37.5) |
| Triglycerides < 150 mg/dl | 357 | 174 (48.7) |
| SBP < 140 mmHg | 643 | 340 (52.9) |
| BMI < 25 kg/m2 | 425 | 126 (29.7) |
| A1c< 7 %, LDL-c < 100 mg/dl y SBP< 140 mmHg | 315 | 14 (4.4) |

IQR: : Interquartile range [25th percentile to 75th percentile]. OAD: Oral antidiabetic drugs (Metformin or glibenclamide). ABI: Ankle-brachial index. A1c: Glycated hemoglobin. SBP: Systolic blood pressure. BMI: Body mass index. a. An alteration in eight assessed areas. IQR b. Delayed or absent response to a 128 Hz tuning fork. c. Slow or absent reflex. d. Estimated glomerular filtration rate calculated by the CKD-EPI

### Prevalence of PAD and Calcification

Peripheral arterial disease (PAD) was present according to the criteria of high, low, and average ABI in 7.8%, 28.2%, and 15.4%, respectively. Arterial calcification was present in 18.2%, 16.2%, and 11% for the same criteria (**Table 2**).

**Table 2.** Prevalence of PAD and calcification according to ABI criteria in patients with diabetes mellitus treated at Hospital María Auxiliadora 2015-2020.

|  | Highest ABI |  | Average ABI |  | Lowest ABI |  |
| --- | --- | --- | --- | --- | --- | --- |
|  | criteria |  | criteria |  | criteria |  |
|  | N | Prevalence<br>(95% CI) | N | Prevalence<br>(95% CI) | N | Prevalence<br>(95% CI) |
| **All** |  |  |  |  |  |  |
| PAD | 50 | 7.8 (5.8-10.1) | 99 | 15.4 (12.7-18.4) | 181 | 28.2 (24.7-31.7) |
| Normal | 476 | 74.0 (70.5-77.3) | 473 | 73.6 (69.9-76.9) | 358 | 55.7 (51.7-59-.5) |
| Arterial calcification | 117 | 18.2 (15.3-21.4) | 71 | 11.0 (8.7-13.7) | 104 | 16.2 (13.4-19.3) |
| **Male** |  |  |  |  |  |  |
| PAD | 16 | 8.3 (4.8-13.0) | 28 | 14.4 (9.8-20.2) | 56 | 28.9 (22.6-35.7) |
| Normal | 141 | 72.7 (65.8-78.8) | 143 | 73.7 (66.9-79.8) | 105 | 54.1 (46.8-61.3) |
| Arterial calcification | 37 | 19.1 (13.8-25.3) | 23 | 11.9 (7.7-17.2) | 34 | 17.0 (12.5-23.6) |
| **Female** |  |  |  |  |  |  |
| PAD | 34 | 7.6 (5.3-10.4) | 71 | 15.8 (12.5-19.6) | 125 | 27.8 (23.7-32.3) |
| Normal | 335 | 74.6 (70.3-78.6) | 330 | 73.5 (69.1-77.5) | 253 | 56.4 (51.6-60.9) |
| Arterial calcification | 80 | 17.8 (14.3-21.7) | 48 | 10.7 (7.9-13.9) | 71 | 15.8 (12.6-19.5) |

ABI: Arm Brachial Index. PAD: Peripheral arterial disease

Normal ABI values were observed in 74% of cases using the highest pressure, 73.6% with the average pressure, and 55.7% with the lowest pressure.

**Supplementary Tables S1, S2, and S3** detail the absolute and relative frequencies of each clinical-demographic variable according to the ABI results for each of the three methods. **Supplementary Table S4** describes the means of each quantitative variable according to the ABI results.

### Characteristics Associated with PAD

For PAD, age was the only factor associated across all three methods. For the high ABI method, in subjects over 75 years old, PAD increased by 7.1 times compared to those under 60 years old (aPR 8.2; 95% CI 1.11 – 60.1). Patients with a history of ulcers showed nearly three times higher prevalence of PAD (aPR 2.88; 95% CI 1.11 – 7.49; P < 0.05). Conversely, the prevalence of PAD decreased by 88% in obese patients compared to those of normal weight (aPR 0.11; 95% CI 0.03 -0.49; p < 0.05). For the low ABI method, a significant association with older age was also observed; in those over 75 years, PAD increased by 81% compared to those under 60 years (RP 1.81; 95% CI 1.11 – 2.95), in conjunction with patients with chronic kidney disease where the prevalence of PAD was 55% (aPR 1.55; 95% CI 1.08 - 2.23; p < 0.05). For the average ABI, the prevalence increased by 1.87 times (aPR 2.87; 95% CI 1.28 – 6.43), and the presence of hypertension increased the prevalence of PAD by 87% compared to those without this history (aPR 1.87; 95% CI 1.12 - 3.13; p < 0.05) (**Table 3**).

**Table 3.** Prevalence Ratio of Peripheral arterial disease vs. Normal according to Clinical-Demographic Characteristics of Patients with Diabetes Mellitus Treated at Hospital María Auxiliadora 2015-2020.

|  | Highest ABI criteria |  |  |  | Lowest ABI criteria |  |  |  | Average ABI criteria |  |  |  |
| --- | --- | --- | --- | --- | --- | --- | --- | --- | --- | --- | --- | --- |
|  | Crude |  | Adjusted |  | Crude |  | Adjusted |  | Crude |  | Adjusted |  |
|  | PR (95% CI) | p-value | PR (95% CI) | p-value | PR (95% CI) | p-value | PR (95% CI) | p-value | PR (95% CI) | p-value | PR (95% CI) | p-value |
| **Genre** |  |  |  |  |  |  |  |  |  |  |  |  |
| Male | 1.00 |  |  |  | 1.00 |  |  |  | 1.00 |  |  |  |
| Female | 0.90 (0,51-1,59) | 0.726 |  |  | 0.95 (0,73 -1.22) | 0.699 |  |  | 1.08 (0.72-1.61) | 0.701 |  |  |
| **Age (years),** |  |  |  |  |  |  |  |  |  |  |  |  |
| Under 60 | 1.00 |  | 1.00 |  | 1.00 |  | 1.00 |  | 1.00 |  | 1.00 |  |
| 60 to 74.9 | 5.91 (2.35-14.8) | <0.001 | 10.4 (1.59-68.3) | **0.014** | 1.31 (0.99 -1.71) | 0.051 | 1.26 (0.81-1.97) | 0.295 | 2.47 (1.57-3.91) | 0.000 | 2.35 (1.71-4.73) | **0.016** |
| 75 or older. | 7.39 (2.66 – 20.5) | <0.001 | 8.20 (1.11-60.1) | **0.038** | 1.64 (1.17-2.32) | 0.004 | 1.81 (1.11-2.95) | **0.017** | 2.72 (1.53-4.83) | 0.001 | 2.87 (1.28-6.43) | **0.010** |
| **Instrucción** |  |  |  |  |  |  |  |  |  |  |  |  |
| Illiterate | 1.00 |  |  |  | 1.00 |  |  |  | 1.00 |  |  |  |
| Elementary | 0.83 (0.34- 2.06) | 0.699 |  |  | 0.85 (0.56-1.31) | 0.476 |  |  | 0.52 (0.30-0.91) | 0.023 |  |  |
| High-School | 0.59 (0.23-1.46) | 0.253 |  |  | 0.76 (0.49 -1.15) | 0.195 |  |  | 0.53 (0.31-0.90) | 0.020 |  |  |
| College | 0.24 (0.5-1.20) | 0.083 |  |  | 0.53 (0.29 -0.98) | 0.044 |  |  | 0.29 (0.12-0.72) | 0.008 |  |  |
| **Tiempo diabetes** |  |  |  |  |  |  |  |  |  |  |  |  |
| Under 10 | 1.00 |  | 1.00 |  | 1.00 |  | 1.00 |  | 1.00 |  | 1.00 |  |
| 10 to 19.9 | 1.62 (0.87-3.01) | 0.128 | 0.82 (0.35-1.92) | 0.653 | 1.33 (1.02-1.73) | 0.035 | 1.24 (0.83-1.86) | 0.287 | 1.42 (0.94-2.15) | 0.092 | 1.12 (0.61-2.07) | 0.702 |
| 20 or more | 3.67 (1.96-6.90) | <0.001 | 1.57 (0.62-3.94) | 0.337 | 1.89 (1.40 -2.56) | <0.001 | 1.39 (0.85-2.26) | 0.179 | 2.22 (1.42-3.47) | <0.001 | 1.46 (0.74-2.85) | 0.270 |
| **Tratamiento de diabetes** |  |  |  |  |  |  |  |  |  |  |  |  |
| Not medication | 1.00 |  |  |  | 1.00 |  |  |  | 1.00 |  |  |  |
| OAD | 0.58 (0.22-1.52) | 0.268 |  |  | 0.71 (0.42-1.21) | 0.213 |  |  | 0.66 (0.31-1.40) | 0.284 |  |  |
| Inuline & OAD | 0.48 (0.09-2.41) | 0.374 |  |  | 0.62 (0.28 -1.41) | 0.259 |  |  | 0.81 (0.29-2.25) | 0.697 |  |  |
| Only Insuline | 0.65 (0.23-1.79) | 0.409 |  |  | 1.17 (0.70-1.95) | 0.544 |  |  | 0.94 (0.44-2.01) | 0.881 |  |  |
| **Altered foot pulses <sup>a</sup>** |  |  |  |  |  |  |  |  |  |  |  |  |
| No | 1.00 |  |  |  | 1.00 |  |  |  | 1.00 |  |  |  |
| Yes | 3.16 (1.88-5.32) | <0.001 |  |  | 3.00 (2.41- 3.73) | <0.001 |  |  | 3.63 (2.56-5.14) | <0.001 |  |  |
| **Peripheral neuropathy<sup>b</sup>** |  |  |  |  |  |  |  |  |  |  |  |  |
| No | 1.00 |  | 1.00 |  | 1.00 |  | 1.00 |  | 1.00 |  | 1.00 |  |
| Yes | 1.74 (1.02-2.97) | 0.039 | 1.31 (0.56-3.06) | 0.531 | 1.29 (1.02 – 1.66) | 0.035 | 1.10 (0.74-1.64) | 0.613 | 1.41 (0.97-2.03) | 0.067 | 1.06 (0.61-1.86) | 0.827 |
| **Ulcer history** |  |  |  |  |  |  |  |  |  |  |  |  |
| No | 1.00 |  | 1.00 |  | 1.00 |  | 1.00 |  | 1.00 |  | 1.00 |  |
| Yes | 3.30 (1.79 – 6.06) | <0.001 | 2.88 (1.11-7.49) | **0.030** | 1.63 (1.18 -2.25) | 0.003 | 1.51 (0.89-2.55) | 0.119 | 1.57 (0.93-2.66) | 0.088 | 1.71 (0.82-3.56) | 0.149 |
| **High blood pressure history** |  |  |  |  |  |  |  |  |  |  |  |  |
| No | 1.00 |  | 1.00 |  | 1.00 |  | 1.00 |  | 1.00 |  | 1.00 |  |
| Yes | 1.89 (1.01 – 3.54) | 0.044 | 1.62 (0.81-3.23) | 0.168 | 1.29 (0.96 – 1.75) | 0.090 | 1.36 (0.96-1.93) | 0.088 | 1.96 (1.26-3.05) | 0.003 | 1.87 (1.12-3.13) | **0.016** |
| **BMI <sup>c</sup>** |  |  |  |  |  |  |  |  |  |  |  |  |
| Under 25 kg/m<sup>2</sup> | 1.00 |  | 1.00 |  | 1.00 |  | 1.00 |  | 1.00 |  | 1.00 |  |
| 25 to 29.9 kg/m<sup>2</sup> | 0.51 (0.26 – 0.97) | 0.041 | 0.36 (0.17-0.76) | **0.008** | 0.94 (0.68 – 1.28) | 0.706 | 0.81 (0.56-1.18) | 0.282 | 0.66 (0.41-1.05) | 0.082 | 0.57 (0.33-0.97) | **0.041** |
| 30 kg/m<sup>2</sup> or more | 0.11 (0.03 – 0.49) | <0.001 | 0.11 (0.01 – 0.87) | **0.037** | 0.63 (0.41 – 0.97) | 0.036 | 0.81 (0.51-1.30) | 0.509 | 0.45 (0.23-0.85) | 0.014 | 0.49 (0.23-1.04) | 0.065 |
| **Kidney disease by CKDEPI <sup>d</sup>** |  |  |  |  |  |  |  |  |  |  |  |  |
| No | 1.00 |  | 1.00 |  | 1.00 |  | 1.00 |  | 1.00 |  | 1.00 |  |
| Yes | 2.27 (1.13-4.54) | 0.020 | 1.57 (0.85-2.91) | 0.145 | 1.79 (1.32 -2.43) | <0.001 | 1.55 (1.08-2.23) | **0.015** | 1.91 (1.15-3.17) | 0.012 | 1.43 (0.83-2.49) | 0.194 |
| **A1C** |  |  |  |  |  |  |  |  |  |  |  |  |
| < 7% | 1.00 |  |  |  | 1.00 |  |  |  | 1.00 |  |  |  |
| ≥ 7% | 1.19 (0.45-3.08) | 0.709 |  |  | 1.15 (0.74 – 1.79) | 0.516 |  |  | 2.02 (0.89-4.58) | 0.090 |  |  |

PAD: Peripheral arterial disease. ABI: Ankle-brachial index. OAD: Oral antidiabetic drugs (Metformin or glibenclamide). a. Positive if there is an absence of pulse in any of the arteries: right or left pedal, right or left
posterior tibial. b. Positive if the monofilament or tuning fork test is altered. c. Body mass index: Normal weight (BMI < 25), Overweight (BMI 25 to 29.9), Obesity (BMI: 30 and above). d. Chronic kidney disease. (Yes:
Estimated glomerular filtration rate calculated by the CKD-EPI formula less than 60 ml/min)

### Characteristics Associated with Arterial Calcification (AC)

In arterial calcification (AC), diabetes duration was the only factor that was associated across all three methods of analysis. In the high ABI method, individuals with more than 20 years of diabetes had an AC prevalence 2.79 times higher compared to those with less than 10 years (aPR 3.79; 95% CI 2.29 – 6.25); in the average ABI, the prevalence increased by 2.12 times (aPR 3.12; 95% CI 1.54 - 6.30; p < 0.001), and for the low ABI, it increased by 2.01 times (aPR 3.01; 95% CI 1.88 - 4.82; p < 0.001).

Regarding elevated BMI (obesity), in the average ABI, the prevalence of AC more than doubled compared to subjects with normal weight (aPR 2.26; 95% CI 1.01 - 5.04; p < 0.05). Similarly, it increased in the low ABI (aPR 2.05; 95% CI 1.21 - 3.45; p < 0.05). For the high ABI, there was a 38% increase, although it was not significant (aPR 1.38; 95% CI 0.76 - 2.50; p=0.287).

In patients with a history of ulcers, only in the low ABI, there was double the prevalence of AC compared to those without this history (aPR 2.01, 95% CI 1.24 - 3.25, p < 0.01). In the other two methods, there was only a 40% increase, which was not significant (**Table 4**).

**Table 4.**
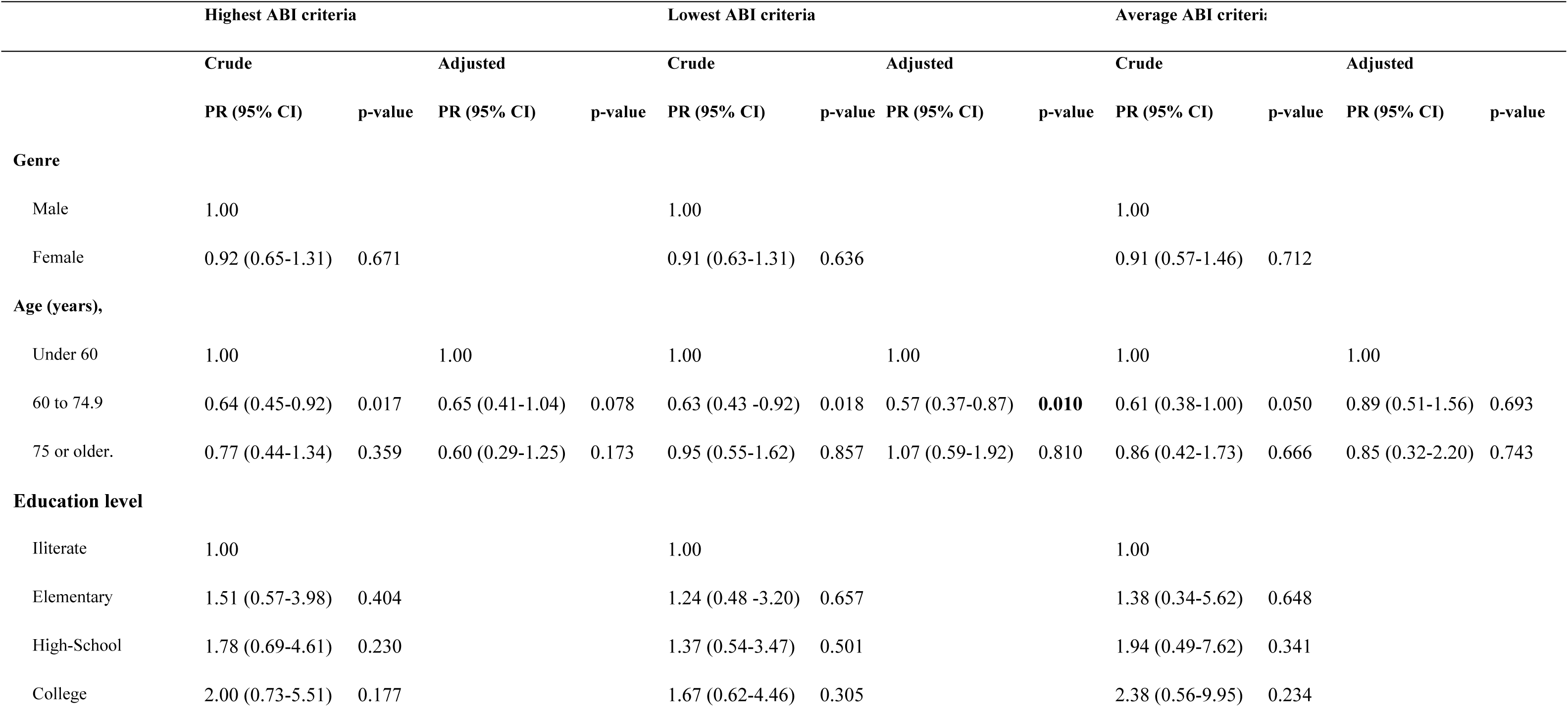
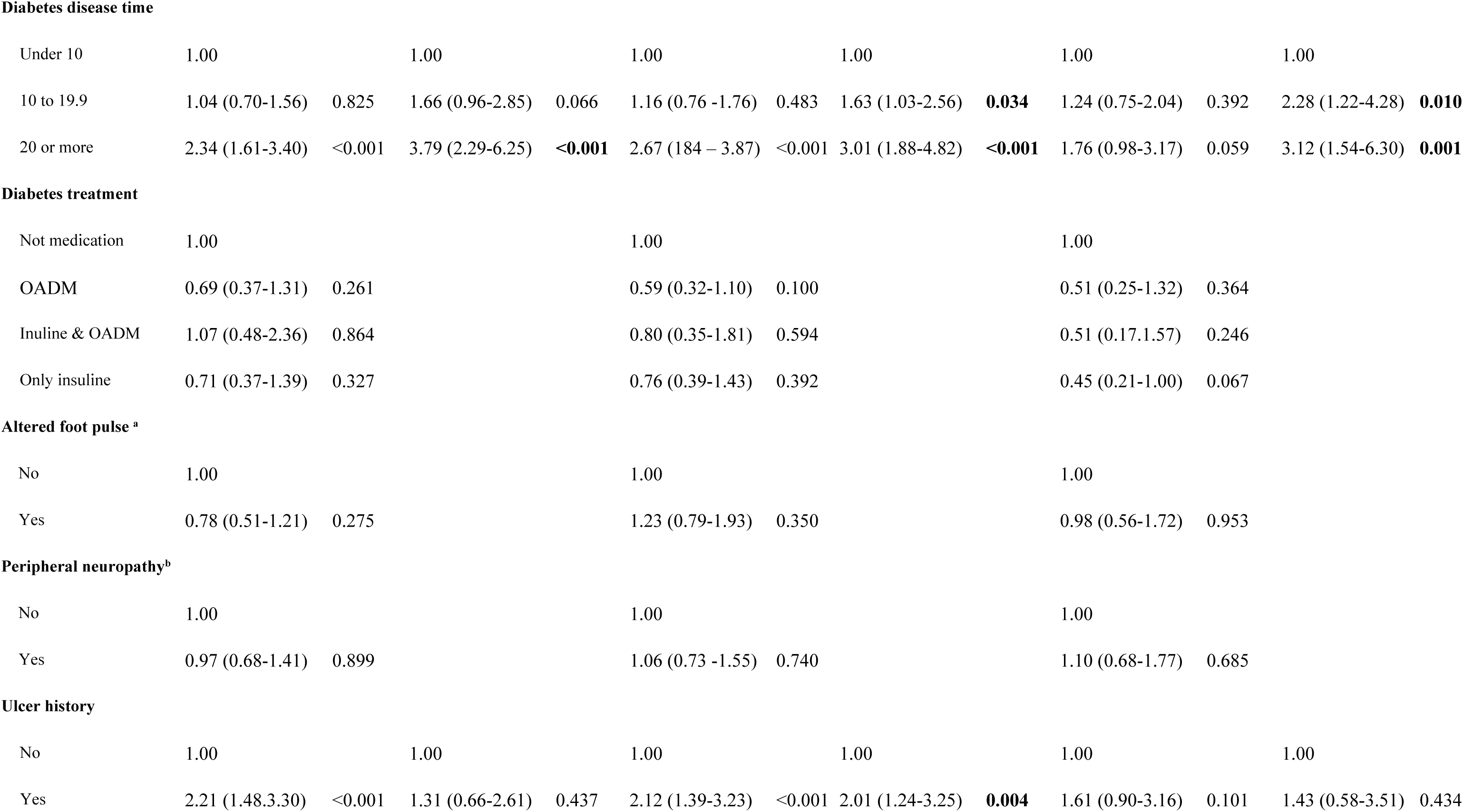
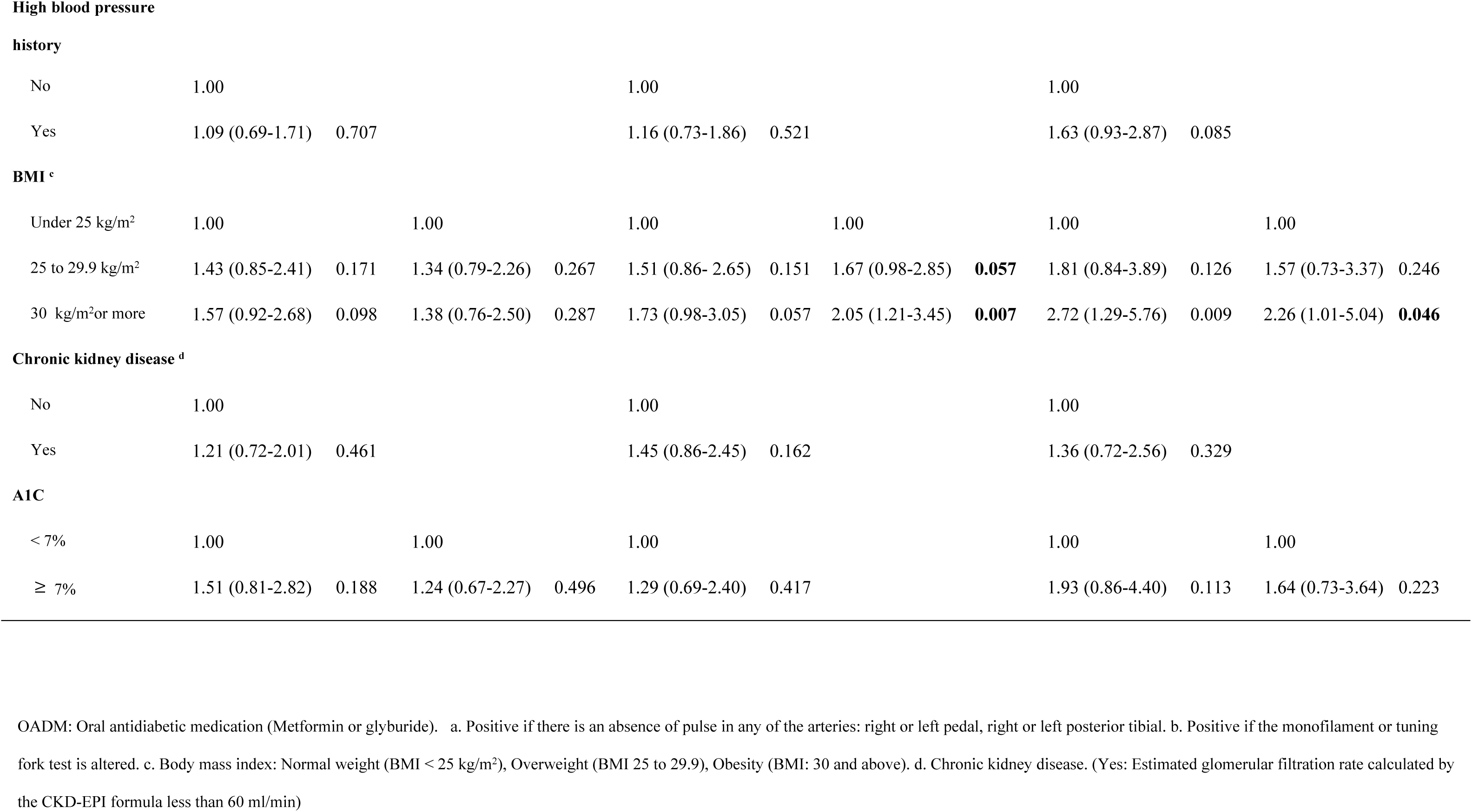
Prevalence Ratio of arterial calcification vs. Normal according to Clinical-Demographic Characteristics of Patients with Diabetes Mellitus Treated at Hospital María Auxiliadora 2015-2020.

|  | Highest ABI criteria |  |  |  | Lowest ABI criteria |  |  |  | Average ABI criteria |  |  |  |
| --- | --- | --- | --- | --- | --- | --- | --- | --- | --- | --- | --- | --- |
|  | Crude |  | Adjusted |  | Crude |  | Adjusted |  | Crude |  | Adjusted |  |
|  | PR (95% CI) | p-value | PR (95% CI) | p-value | PR (95% CI) | p-value | PR (95% CI) | p-value | PR (95% CI) | p-value | PR (95% CI) | p-value |
| **Genre** |  |  |  |  |  |  |  |  |  |  |  |  |
| Male | 1.00 |  |  |  | 1.00 |  |  |  | 1.00 |  |  |  |
| Female | 0.92 (0.65-1.31) | 0.671 |  |  | 0.91 (0.63-1.31) | 0.636 |  |  | 0.91 (0.57-1.46) | 0.712 |  |  |
| **Age (years),** |  |  |  |  |  |  |  |  |  |  |  |  |
| Under 60 | 1.00 |  | 1.00 |  | 1.00 |  | 1.00 |  | 1.00 |  | 1.00 |  |
| 60 to 74.9 | 0.64 (0.45-0.92) | 0.017 | 0.65 (0.41-1.04) | 0.078 | 0.63 (0.43 -0.92) | 0.018 | 0.57 (0.37-0.87) | **0.010** | 0.61 (0.38-1.00) | 0.050 | 0.89 (0.51-1.56) | 0.693 |
| 75 or older. | 0.77 (0.44-1.34) | 0.359 | 0.60 (0.29-1.25) | 0.173 | 0.95 (0.55-1.62) | 0.857 | 1.07 (0.59-1.92) | 0.810 | 0.86 (0.42-1.73) | 0.666 | 0.85 (0.32-2.20) | 0.743 |
| **Education level** |  |  |  |  |  |  |  |  |  |  |  |  |
| Illiterate | 1.00 |  |  |  | 1.00 |  |  |  | 1.00 |  |  |  |
| Elementary | 1.51 (0.57-3.98) | 0.404 |  |  | 1.24 (0.48 -3.20) | 0.657 |  |  | 1.38 (0.34-5.62) | 0.648 |  |  |
| High-School | 1.78 (0.69-4.61) | 0.230 |  |  | 1.37 (0.54-3.47) | 0.501 |  |  | 1.94 (0.49-7.62) | 0.341 |  |  |
| College | 2.00 (0.73-5.51) | 0.177 |  |  | 1.67 (0.62-4.46) | 0.305 |  |  | 2.38 (0.56-9.95) | 0.234 |  |  |
| **Diabetes disease time** |  |  |  |  |  |  |  |  |  |  |  |  |
| Under 10 | 1.00 |  | 1.00 |  | 1.00 |  | 1.00 |  | 1.00 |  | 1.00 |  |
| 10 to 19.9 | 1.04 (0.70-1.56) | 0.825 | 1.66 (0.96-2.85) | 0.066 | 1.16 (0.76 -1.76) | 0.483 | 1.63 (1.03-2.56) | **0.034** | 1.24 (0.75-2.04) | 0.392 | 2.28 (1.22-4.28) | **0.010** |
| 20 or more | 2.34 (1.61-3.40) | <0.001 | 3.79 (2.29-6.25) | **<0.001** | 2.67 (1.84 – 3.87) | <0.001 | 3.01 (1.88-4.82) | **<0.001** | 1.76 (0.98-3.17) | 0.059 | 3.12 (1.54-6.30) | **0.001** |

**Diabetes treatment**
|  |  |  |  |  |  |  |  |  |  |  |
| --- | --- | --- | --- | --- | --- | --- | --- | --- | --- | --- |
| Not medication | 1.00 |  |  |  | 1.00 |  |  |  | 1.00 |  |
| OADM | 0.69 (0.37-1.31) | 0.261 |  |  | 0.59 (0.32-1.10) | 0.100 |  |  | 0.51 (0.25-1.32) | 0.364 |
| Inuline & OADM | 1.07 (0.48-2.36) | 0.864 |  |  | 0.80 (0.35-1.81) | 0.594 |  |  | 0.51 (0.17-1.57) | 0.246 |
| Only insuline | 0.71 (0.37-1.39) | 0.327 |  |  | 0.76 (0.39-1.43) | 0.392 |  |  | 0.45 (0.21-1.00) | 0.067 |

**Altered foot pulse <sup>a</sup>**
|  |  |  |  |  |  |  |  |  |  |  |
| --- | --- | --- | --- | --- | --- | --- | --- | --- | --- | --- |
| No | 1.00 |  |  |  | 1.00 |  |  |  | 1.00 |  |
| Yes | 0.78 (0.51-1.21) | 0.275 |  |  | 1.23 (0.79-1.93) | 0.350 |  |  | 0.98 (0.56-1.72) | 0.953 |

**Peripheral neuropathy<sup>b</sup>**
|  |  |  |  |  |  |  |  |  |  |  |
| --- | --- | --- | --- | --- | --- | --- | --- | --- | --- | --- |
| No | 1.00 |  |  |  | 1.00 |  |  |  | 1.00 |  |
| Yes | 0.97 (0.68-1.41) | 0.899 |  |  | 1.06 (0.73 -1.55) | 0.740 |  |  | 1.10 (0.68-1.77) | 0.685 |
| **Ulcer history** |  |  |  |  |  |  |  |  |  |  |  |  |
| No | 1.00 |  | 1.00 |  | 1.00 |  | 1.00 |  | 1.00 |  | 1.00 |  |
| Yes | 2.21 (1.48-3.30) | <0.001 | 1.31 (0.66-2.61) | 0.437 | 2.12 (1.39-3.23) | <0.001 | 2.01 (1.24-3.25) | **0.004** | 1.61 (0.90-3.16) | 0.101 | 1.43 (0.58-3.51) | 0.434 |
| **High blood pressure** |  |  |  |  |  |  |  |  |  |  |  |  |
| **history** |  |  |  |  |  |  |  |  |  |  |  |  |
| No | 1.00 |  |  |  | 1.00 |  |  |  | 1.00 |  |  |  |
| Yes | 1.09 (0.69-1.71) | 0.707 |  |  | 1.16 (0.73-1.86) | 0.521 |  |  | 1.63 (0.93-2.87) | 0.085 |  |  |
| **BMI <sup>c</sup>** |  |  |  |  |  |  |  |  |  |  |  |  |
| Under 25 kg/m<sup>2</sup> | 1.00 |  | 1.00 |  | 1.00 |  | 1.00 |  | 1.00 |  | 1.00 |  |
| 25 to 29.9 kg/m<sup>2</sup> | 1.43 (0.85-2.41) | 0.171 | 1.34 (0.79-2.26) | 0.267 | 1.51 (0.86- 2.65) | 0.151 | 1.67 (0.98-2.85) | **0.057** | 1.81 (0.84-3.89) | 0.126 | 1.57 (0.73-3.37) | 0.246 |
| 30 kg/m<sup>2</sup>or more | 1.57 (0.92-2.68) | 0.098 | 1.38 (0.76-2.50) | 0.287 | 1.73 (0.98-3.05) | 0.057 | 2.05 (1.21-3.45) | **0.007** | 2.72 (1.29-5.76) | 0.009 | 2.26 (1.01-5.04) | **0.046** |
| **Chronic kidney disease <sup>d</sup>** |  |  |  |  |  |  |  |  |  |  |  |  |
| No | 1.00 |  |  |  | 1.00 |  |  |  | 1.00 |  |  |  |
| Yes | 1.21 (0.72-2.01) | 0.461 |  |  | 1.45 (0.86-2.45) | 0.162 |  |  | 1.36 (0.72-2.56) | 0.329 |  |  |
| **A1C** |  |  |  |  |  |  |  |  |  |  |  |  |
| < 7% | 1.00 |  | 1.00 |  | 1.00 |  |  |  | 1.00 |  | 1.00 |  |
| ≥ 7% | 1.51 (0.81-2.82) | 0.188 | 1.24 (0.67-2.27) | 0.496 | 1.29 (0.69-2.40) | 0.417 |  |  | 1.93 (0.86-4.40) | 0.113 | 1.64 (0.73-3.64) | 0.223 |

OADM: Oral antidiabetic medication (Metformin or glyburide). a. Positive if there is an absence of pulse in any of the arteries: right or left pedal, right or left posterior tibial. b. Positive if the monofilament or tuning fork test is altered. c. Body mass index: Normal weight (BMI < 25 kg/m<sup>2</sup>), Overweight (BMI 25 to 29.9), Obesity (BMI: 30 and above). d. Chronic kidney disease. (Yes: Estimated glomerular filtration rate calculated by the CKD-EPI formula less than 60 ml/min)

## DISCUSSION

Our study found a wide range in the frequency of PAD and a narrow interval of arterial calcification across the three methods used to determine the ABI. Additionally, the highest prevalence of PAD was obtained using the lowest ankle pressure criterion. Conversely, arterial calcification was more frequent when the highest ABI criterion was applied.

### Peripheral Arterial Disease

There is strong evidence that an ABI less than 0.9 is associated with up to twice the risk of cardiovascular or all-cause mortality in subjects with diabetes. Furthermore, it is noted for its better specificity than sensitivity when using the classic highest ABI criterion. However, considering different ABI cut-off points depending on the method used could optimize risk identification. Our study’s findings on PAD prevalence using various methods are like other studies. In a US study with a multiethnic population, a prevalence of 3.7% was found with the highest ABI, 5.4% for the average ABI, and 14.6% with the lowest ABI [27]. A study in France found a prevalence of 22%, 23%, and 29% for the three measurement modes in the same order [28]. The same US study showed that using different calculation methods can slightly improve prognostic accuracy [27].

On the other hand, a recent clinical guideline agrees on using the lowest ABI criterion for risk stratification and the highest ABI for severity assessment and post-revascularization follow-up [18]. The lowest ABI increases prevalence and sensitivity for identifying high-risk patients, though it decreases specificity for diagnosing those with early-stage disease. In practice, diagnosing asymptomatic PAD with an ABI below 0.9 would elevate cardiovascular risk to higher categories. Considering the European cardiovascular risk score for dyslipidemia management (SCORE), with every diabetic patient having at least moderate cardiovascular risk, the presence of asymptomatic PAD would elevate to a high or very high level and intensify risk factor treatment [29].

Regarding characteristics associated with PAD, our study found an association with advanced age for any of the three methods, and depending on the method, factors varied. Increased prevalence at advanced ages reflects the natural progression of atherosclerosis and vascular risk in patients chronically exposed to diabetes [30]. We found no association with gender; it has been reported that women present a similar risk as men, contrary to previous assertions, requiring similar importance in screening [31]. Another risk factor is ethnicity, where Afro-descendants have a higher likelihood. In our study, the original data did not contain this information, but it wouldn’t be a limitation given the high degree of mixed ethnicity in our country, unlike other South American countries [32].

### Arterial Calcification

Traditionally, AC was considered a generalized passive process typical of advanced age and metabolic alterations; however, it has recently been discovered as an active and regulated process similar to bone metabolism and thus modifiable. It can occur at the intima or media layer, with both coexisting to different extents [33]. An ABI greater than 1.3 demonstrates the existence of medial or Monckeberg calcification in lower limbs, causing vascular stiffness, possibly with low flow, in an artery that cannot be compressed. It was long considered an artifact, requiring other methods like the toe-brachial index, as finger arteries are less susceptible to AC [34]. However, it has been highlighted as a critical cardiovascular risk marker in the general population [35] and in patients with diabetes [36].

In our study, the frequency of subjects with an ABI greater than 1.3 in any limb was determined to be 18.3%, lower than that reported in China conducted in the general population, using an average ABI method finding a frequency of 24.4%. However, as the diagnostic algorithm usually prioritizes PAD, AC may be underreported in those who coexist. In our study, only 2.2% presented both findings. It’s important to note that the recommended cut-off point in clinical guidelines is 1.3 [24] or 1.4 [18,37]. Where a highest CA cut-off values reduce prevalence, it becomes more specific. One cut-off point can be established based on epidemiological criteria such as patient risk, disease prevalence, or costs and infrastructure to provide treatment to positive results.[38]

The most studied factors associated with AC include diabetes mellitus, chronic kidney disease, advanced age, and increased creatinine [39]. In our study, AC was associated with longer diabetes duration for any method. Age was not significant but showed a trend towards being more frequent in younger people, suggesting that in certain patients, AC begins very early. This is why, with the low ABI method, AC is associated with higher BMI. Chronic kidney disease did not increase AC prevalence, despite being present in a quarter of those with data for its calculation.

Being a generalized process, arterial calcification (AC) can be assessed using other diagnostic strategies, both invasive (arterial angiography, intravascular ultrasound) and non-invasive (carotid intima-media thickness, brachial artery flow-mediated dilation, or coronary calcification score), which are currently used as risk markers [40].

### Importance of Findings

The simultaneous identification of PAD and AC in diabetic patients implies a dual approach to preventing amputations and reducing the risk of cardiovascular events. The use of the highest ankle pressure as the numerator in the ABI calculation is associated with increased specificity, minimizing the risk of overdiagnosis in healthy individuals and preventing unnecessary testing and treatment. However, this approach may lead to a higher rate of false negatives, particularly in patients with arterial calcification.[37,38] Utilizing the lowest ankle pressure would identify more individuals at risk of mortality, both from cardiovascular and other causes. These findings support considering different cut-off points for the ABI, as the traditional method (highest pressure) might overlook individuals with significant risk.[41]

Furthermore, it is necessary to report AC as an additional risk factor in regular screening to identify them early, and not only view it as a technical issue requiring additional imaging techniques to determine PAD. It would also be essential to locally validate its association with hard outcomes such as cardiovascular disease, major amputation or frailty. [42]

### Limitations and Strengths of the Study

The use of a hospital-based study design may limit the generalizability of the findings, as the selected sample does not necessarily represent the entire population of type 2 diabetes mellitus patients in different care settings. This could affect the extrapolation of results to other patient groups outside the hospital setting. Additionally, the initial focus on determining PAD might have led to underreporting of AC, given that if a patient had PAD in one leg and AC in the other, PAD would take precedence. Moreover, nearly 40% of initial records had to be excluded due to incomplete ABIs; however, the number of included subjects provides good precision for estimating prevalences. Similarly, there was insufficient data on past medical history, body mass index, glycated hemoglobin, and chronic kidney disease, so associations may be limited by sample size. Some variables such as duration of diabetes may present as recall bias, and conditions like diabetic retinopathy may be underreported as they came from self-report and not active searches. A strength is that the ABI was performed by personnel trained in diabetic foot care and with a mercury sphygmomanometer, and peripheral neuropathy data were obtained through objective tests.

### Conclusions

Using different ABI methods, we observed a prevalence of PAD ranging from 7.8% to 28%, and a prevalence of CA between 11% and 18% among patients with type 2 diabetes mellitus at a public hospital in Peru. The method with the lowest criterion showed the highest prevalence. It is more frequent with increasing age and longer duration of diabetes for any of the three methods. Further research is necessary to verify if the new ABI calculation methods provide improved accuracy in predicting complications

## Data Availability

All relevant data are within the manuscript and its Supporting Information files.

## Supplementary material

**Tabla S1. Prevalence of PAD and AC based on the highest ABI criterion according to clinical-demographic characteristics.**

**Tabla S2. Prevalence of PAD and AC based on the lowest ABI criterion according to clinical-demographic characteristics**

**Tabla S3. Prevalence PAD and AC based on the average ABI criterion according to clinical-demographic characteristics.**

**Table S4. Demographic and clinical numerical variables of patients with diabetes mellitus**

